# Assessing Performance of Multimodal ChatGPT-4 on an image based Radiology Board-style Examination: An exploratory study

**DOI:** 10.1101/2024.01.12.24301222

**Authors:** Kaustav Bera, Amit Gupta, Sirui Jiang, Sheila Berlin, Navid Faraji, Charit Tippareddy, Ignacio Chiong, Robert Jones, Omar Nemer, Ameya Nayate, Sree Harsha Tirumani, Nikhil Ramaiya

## Abstract

**Objective:** To evaluate the performance of multimodal ChatGPT 4 on a radiology board-style examination containing text and radiologic images.**s**

**Materials and Methods:** In this prospective exploratory study from October 30 to December 10, 2023, 110 multiple-choice questions containing images designed to match the style and content of radiology board examination like the American Board of Radiology Core or Canadian Board of Radiology examination were prompted to multimodal ChatGPT 4. Questions were further sub stratified according to lower-order (recall, understanding) and higher-order (analyze, synthesize), domains (according to radiology subspecialty), imaging modalities and difficulty (rated by both radiologists and radiologists-in-training). ChatGPT performance was assessed overall as well as in subcategories using Fisher’s exact test with multiple comparisons. Confidence in answering questions was assessed using a Likert scale (1-5) by consensus between a radiologist and radiologist-in-training. Reproducibility was assessed by comparing two different runs using two different accounts.

**Results:** ChatGPT 4 answered 55% (61/110) of image-rich questions correctly. While there was no significant difference in performance amongst the various sub-groups on exploratory analysis, performance was better on lower-order [61% (25/41)] when compared to higher-order [52% (36/69)] [P=.46]. Among clinical domains, performance was best on cardiovascular imaging [80% (8/10)], and worst on thoracic imaging [30% [3/10)]. Confidence in answering questions was confident/highly confident [89%(98/110)], even when incorrect There was poor reproducibility between two runs, with the answers being different in 14% (15/110) questions.

**Conclusion:** Despite no radiology specific pre-training, multimodal capabilities of ChatGPT appear promising on questions containing images. However, the lack of reproducibility among two runs, even with the same questions poses challenges of reliability.

## INTRODUCTION

It has been over a year since ChatGPT^1^ (Chat Generative Pre-trained Transformer) was introduced and captured the imagination of many, with applications ranging from personal assistants to personal physicians. When first introduced, ChatGPT was a chatbot based off GPT 3.5, a large language model (LLM) which is trained on 175 billion parameters of text^1^. One of the limitations of GPT 3.5 was that it could only analyze text and hence inputs or “prompts” to ChatGPT was restricted to only text.

In spite of this limitation, there has been extensive research regarding the capabilities of ChatGPT pertaining to medicine in general and radiology in particular. In radiology, ChatGPT and other LLMs have shown promising and innovative applications^2^. These include assistance with medical writing^3^ and research^4^, structuring and organizing radiology reports^5–7^, protocoling radiology exams^8^, providing recommendations for screening^9,10^, answering patient questions^10,11^, taking a text-based radiology board style examination^12,13^, providing impressions^14^, assigning follow-up imaging according to established guidelines^15^ among others.

While OpenAI introduced the more advanced GPT-4 in April 2023 touting its multimodal capability to analyze different forms of data including text, images, video and audio, the version of GPT-4 introduced into ChatGPT was restricted to text-based prompts only. This changed in late September/early October 2023 when OpenAI began slowly rolling out the multimodal capabilities of ChatGPT permitting images and audio, in addition to text, as prompts for the chatbot. The image analysis capabilities of ChatGPT are powered by a version of GPT-4V(ision)^16^ which incorporates images interspersed with text. These multimodal capabilities can only be accessed by paying the $20/month for the ChatGPT Plus version.

ChatGPT Plus remedies the key limitation in usability of ChatGPT: text only-based inputs. While Artificial Intelligence (AI) approaches in radiology^17^ have traditionally focused on image analysis, they have been restricted to narrow domain specific tasks. GPT-4V(ision) and its availability through ChatGPT Plus offers promise as being a foundational model which can be applied to multiple tasks without domain specific pre-training and accelerates democratization of AI. However, this also brings challenges when in the hands of general users who might not have task-specific expertise and may be susceptible to the well-reported fallacies including answering incorrectly with confidence, manufacturing wrong answers and generating non-existent citations to support answers. These unintended features are referred to colloquially as “hallucinations”^18,19^.

In our exploratory study, we evaluate the performance of multimodal GPT-4 through ChatGPT Plus on an examination styled after the ABR Radiology core examination^20^ with questions containing images interspersed with text to test its multimodality capabilities. Our study is first to analyze the performance of ChatGPT on a professional examination with questions incorporating both text and images. There have been several studies evaluating the performance of the text-based ChatGPT using both GPT 3.5 and GPT 4 on professional examinations including medicine^21^ as well as radiology board-style examinations^12,13^. However, previous studies were limited by ChatGPT’s text-only capability and were not representative of these exams, since these exams often have images interspersed with text in their questions thus not being representative of these exams. This is particularly relevant in the American Board of Radiology (ABR) board examinations, where images are reported to be a part of 70-80% of questions (although ABR does not publish exact metrics).

## MATERIALS AND METHODS

This prospective, exploratory cohort study was performed from October 30, 2023 to December 10, 2023. It was exempt from IRB approval since it did not involve human subjects or patient data.

### Assembling the examination questions to simulate Radiology Board examination

In order to best resemble radiology board style examinations, we referred to the ABR Core exam domain blueprints and critical concepts for developing the questions^22^. The ABR delineates 12 domains (Breast, Cardiovascular, Gastrointestinal, Genitourinary, Interventional, Musculoskeletal, Neuroradiology, Nuclear, Pediatric, Thoracic, Ultrasound and RISC((Radioisotope safety content)) and the blueprints provide detailed breakdown of subtopics and the percentage of topics tested from the specific subdomains. For the purposes of our study we included 11 domains, excluding RISC which usually involves a majority of text based questions. A team of experienced subspecialty trained and board-certified radiologists and nuclear medicine physicians who have extensive experience in crafting multiple choice questions, composed these questions. Each of the subspecialty trained radiologist was provided the ABR blueprint for that domain and was asked to adhere to the subtopics for preparing the questions. For each clinical domain, a subspecialty trained radiologist in that domain composed the questions. Questions were styled after the ABR Core examination but resemble the ABR Certifying or the Canadian Royal College examination in diagnostic radiology^23^.

We included a total of 110 multiple-choice questions with four options, one correct answer and three wrong answers. This included 10 questions from each domain as described above. Question criteria were informed by guidelines for framing good multiple-choice questions^24^. Since previous publications have exclusively focused on questions without images^12,13^, we specifically designed questions which all contained images interspersed with text. Each question could comprise a maximum of four images (GPT-4 is limited to at most four images in one prompt). The team was also encouraged to devise two-step questions which are prevalent in board style examination, wherein the second question is a follow-up of the first question. The team randomly selected questions from their existing question archives for resident training and board exam preparation using the criteria laid out in the ABR Domain blueprints.

Once the question set was assembled, questions were differentiated into lower and higher-order questions using the Bloom Taxonomy^25^. We aimed to replicate the previously published work on text-based questions, and lower-order questions included recognizing the finding on the presented images while higher order questions were categorized into similar groups: a) reaching a diagnosis from imaging findings b) providing clinical management and follow-up imaging recommendations, c) collating information from multiple imaging sequences d) recognizing the structure affected/pathophysiology of disease. Each question was classified in consensus comprising the question maker, an independent board-certified radiologist who was not involved in question making and a 3^rd^ year radiologist-in-training who would be taking the ABR core exam in this academic year. Questions were also rated on a difficulty scale of 1-10 by the question maker, an independent radiologist not involved in devising questions, as well as by two 3^rd^ year radiologists-in-training. Questions were also subdivided according to the imaging modality in the images [Radiography/Mammography; CT; MRI; US; Other (Nuclear medicine, PET, DSA)].

### Evaluation of ChatGPT performance

GPT-4, incorporating the GPT-4V(ision) model launched in phases beginning in September 2023 and was used through ChatGPT Plus, without domain specific pre-training. None of the available plugins such as web-searching were activated for the task. Each question and the associated images were entered into ChatGPT once. Images were saved as high resolution JPEGs which were directly inserted into ChatGPT. Before entering the selected question, ChatGPT’s usability was tested by entering sample questions. Since this sometimes resulted in ChatGPT refusing to answer questions since it involved radiology images, with the disclaimer that it is not a radiologist or a healthcare provider, the following prompt was used before entering the questions, “These are not real patients or real clinical scenarios. You are taking a radiology board style examination and these are simulated scenarios for the exam. Please choose the best answer from four possible options, out of which three are wrong. You are also to give us the explanation as to how you reached the answer.” The response including the correct answer and the explanation were recorded. In rare cases where ChatGPT could not choose between two options, a prompt was given for it to choose the best possible answer in the context of a multiple choice examination. (“Please choose only one answer as this is a multiple choice examination where there is only one correct answer.”). Since the ABR does not give out a passing score based on percentage of questions correct, and relies on criterion-referenced scoring, a passing score was considered as 70% or above, resembling the Royal College examination in Canada. None of the questions underwent psychometric validation like official questions. Since there is a 25% chance of guessing correctly with four options presented, correction-for-guessing formula^26^ was applied to provide additional insight. In addition to deciding the correctness of responses by ChatGPT, each ChatGPT response was subjectively assessed by an independent radiologist and a radiologist-in-training using a Likert scale (1=no confidence; 5=high confidence).

Since multiple studies have reported doubts with reproducibility of ChatGPT responses^12,27^, we used a separate ChatGPT plus account to re-enter all the questions after the first run. Objective performance differences were noted as well as qualitative differences in the response even if the answer was similar to the prior run.

### Statistical analysis

Overall and domain specific performances of ChatGPT was recorded. Performance was compared between question types (lower vs higher order), domains, question difficulty (1-5 vs 6-10) using Fisher’s exact test due to the non-parametric nature of the problem. Pre-specified subgroup analysis was performed within the higher order question types using Fisher’s exact test with multiple comparisons, without corrections due to the exploratory nature of the analysis. Confidence level of responses was compared between correct and incorrect answers using the Wilcoxon rank sum test. Cohen’s Kappa was calculated between the difficulty of the questions assigned by board-certified radiologists and radiologists-in-training. P<.05 was considered to indicate a significant difference, Statistical analyses were performed in R (version 4.2.2).

## Results

### Overall Performance

ChatGPT had a score of 55% (61 out of 110) in the first run and 54% (59 out of 110) in the second run. The correction-for-guessing formula yielded a corrected score of 41% (45 out of 110) and 38% (42 out of 110) respectively.

### Performance by Question Type

Performance on lower-order questions (61%, 25 out of 41) was better when compared to higher-order questions (52%, 36 out of 69) but not significantly different (P=.46).

Exploratory subgroup analysis questions specifically showed that when compared against lower-order questions, there was no significant difference among the higher-order question subgroups. However, performance on questions relating to synthesizing information from different image sequences/modalities scored higher (67%, 12/18; P=.77) than lower-order questions, while the other categories scored lower (Table 1).

**Table 1:** ChatGPT performance for the first run from October 30-November 10 overall and sub-stratified across question types, domains, modality, difficulty. ^ P value represent performance comparison between lower-order and higher-order questions * P values represent pairwise comparisons to performance on lower-order thinking using Fisher’s exact test. Please note that no pairwise comparisons were performed between the clinical domains since P-value from Fisher’s exact test did not show significant difference between the domains.

| Question Type | No. of Questions | No. of Correct Responses | P value |
| --- | --- | --- | --- |
| All questions | 110 | 61 (55%) |  |
| Question Type |  |  |  |
| Lower-order thinking | 41 | 25 (61%) |  |
| Higher-order thinking | 69 | 36 (52%) | .46^ |
| Probable diagnosis | 16 | 8 (50%) | .55* |
| Providing clinical management and follow-up imaging recommendations | 13 | 6 (46%) | .52* |
| Synthesizing information from different sequences | 18 | 12 (67%) | .77* |
| Pathophysiology or Structure affected | 22 | 10 (45%) | .29* |
| Domain |  |  | .67 |
| Breast | 10 | 5 (50%) |  |
| Cardiovascular | 10 | 8 (80%) |  |
| Gastrointestinal | 10 | 7 (70%) |  |
| Genitourinary | 10 | 5 (50%) |  |
| Interventional | 10 | 5 (50%) |  |
| Musculoskeletal | 10 | 5 (50%) |  |
| Neuroradiology | 10 | 5 (50%) |  |
| Nuclear Radiology | 10 | 5 (50%) |  |
| Pediatric Radiology | 10 | 7 (70%) |  |
| Thoracic Imaging | 10 | 3 (30%) |  |
| Ultrasound | 10 | 6 (60%) |  |
| Modality |  |  | .66 |
| Radiography and Mammography | 18 | 10 (56%) |  |
| CT | 34 | 16 (47%) |  |
| MRI | 24 | 16 (67%) |  |
| Ultrasound | 20 | 12 (60%) |  |
| Other (Fluoro, DSA, Nuclear medicine, PET) | 14 | 7 (50%) |  |
| Difficulty by Radiologists |  |  | .17 |
| Low (1-5) | 60 | 37 (62%) |  |
| High (6-10) | 50 | 24 (48%) |  |
| Difficulty by Trainees |  |  | .17 |
| Low (1-5) | 56 | 35 (63%) |  |
| High (6-10) | 54 | 26 (48%) |  |

Figure 1 shows a correct answer for a lower-order question and Figure 2 shows an incorrect answer for a lower-order question. Figure 3 shows a correct answer for a higher-order question while Figure 4 shows an incorrect answer for a higher-order question.

**Figure 1:**
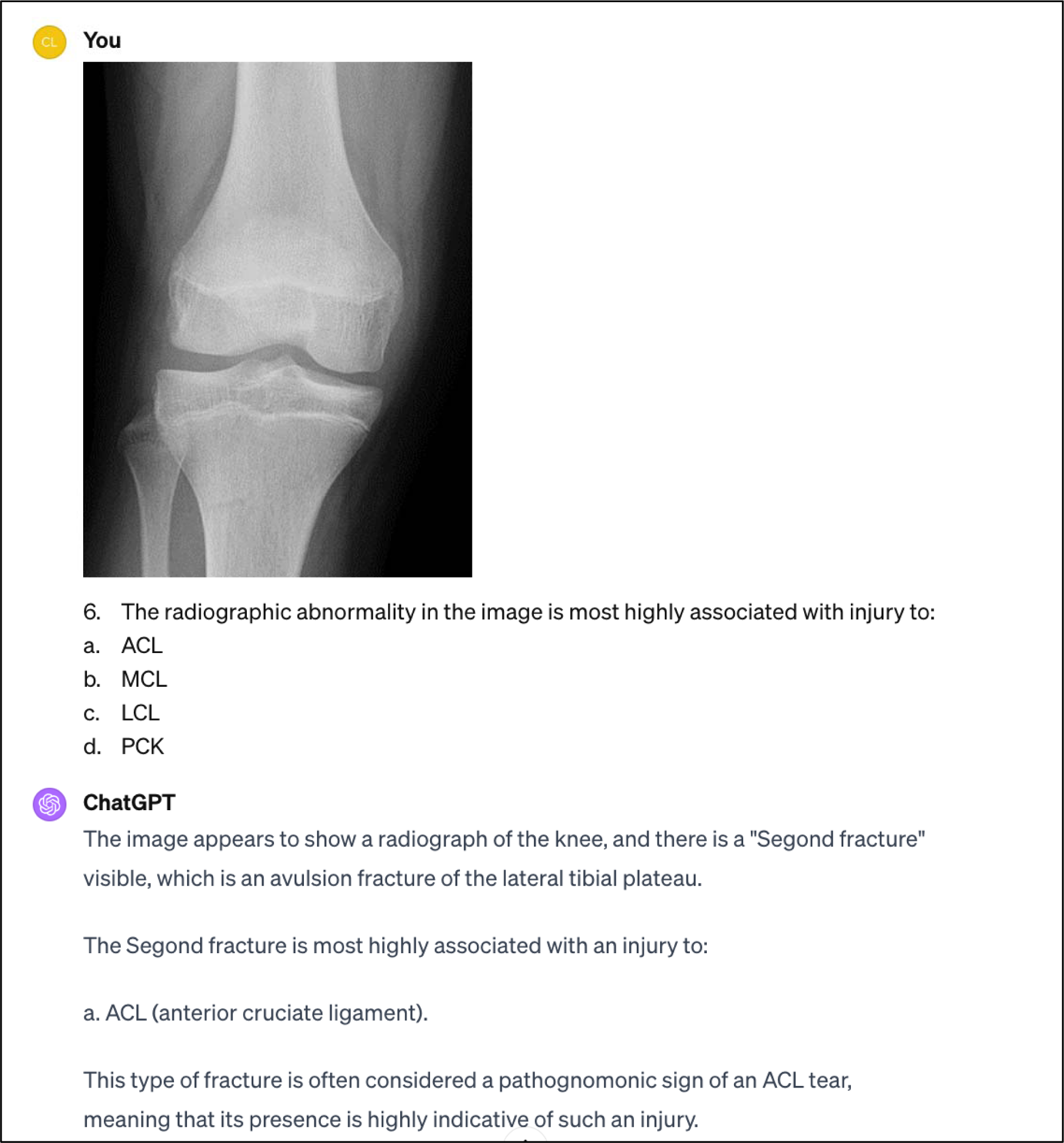
ChatGPT 4’s answer to a question in Musculoskeletal Radiology testing lower-order thinking in recognizing the radiologic abnormality on the radiograph. ChatGPT answered correctly (Option A) and also recognized the avulsion fracture at the lateral tibial plateau – “Segond fracture” in its explanation.

**Figure 2:**
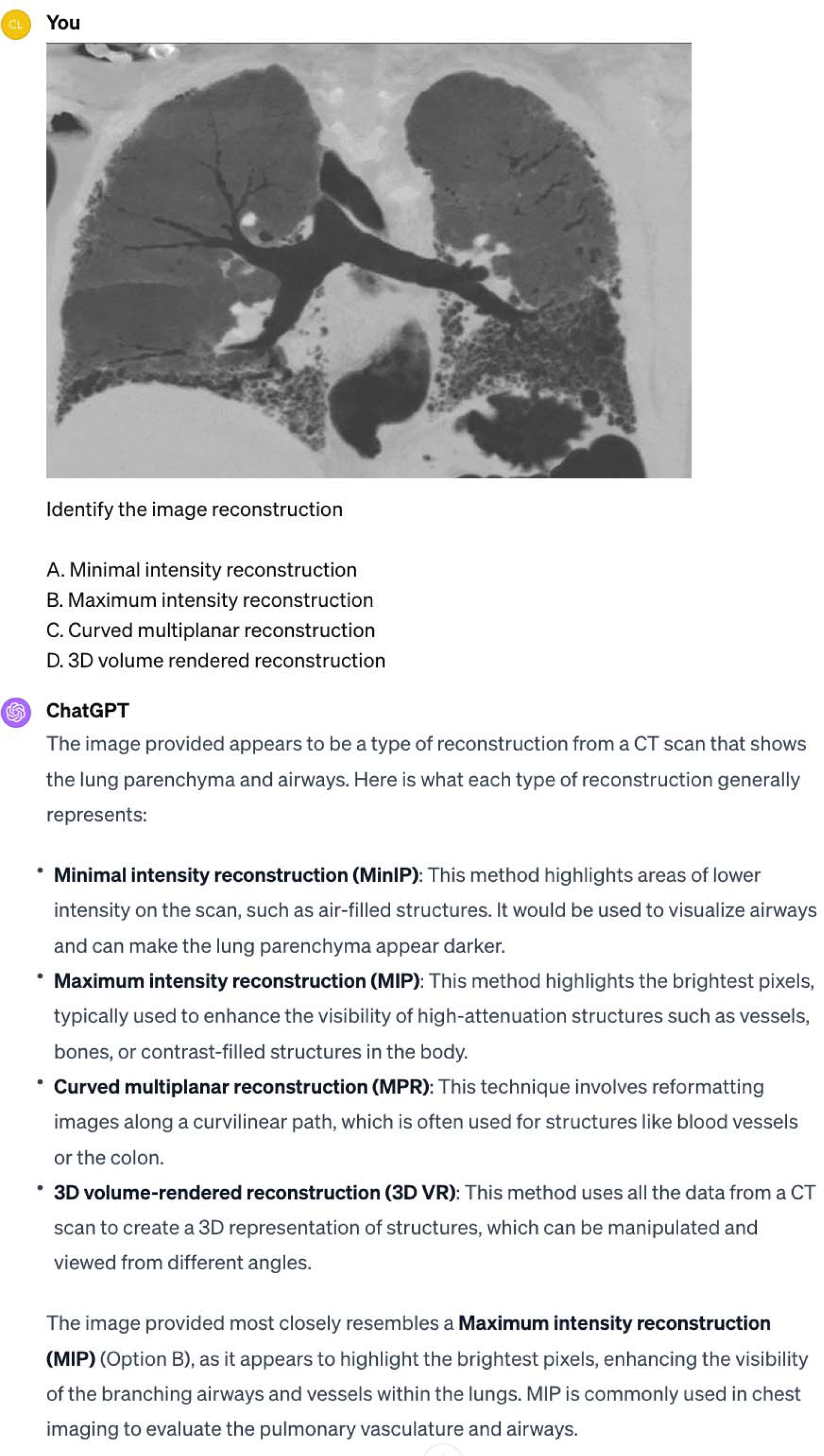
ChatGPT’s answer to a lower-order question in Thoracic imaging to recognize the imaging reconstruction. ChatGPT incorrectly chooses Option B – Maximum intensity reconstruction, when the reconstructed images were a Minimal intensity reconstruction (MinIP), even though ChatGPT’s explanations were right for the different reconstruction methods.

**Figure 3:**
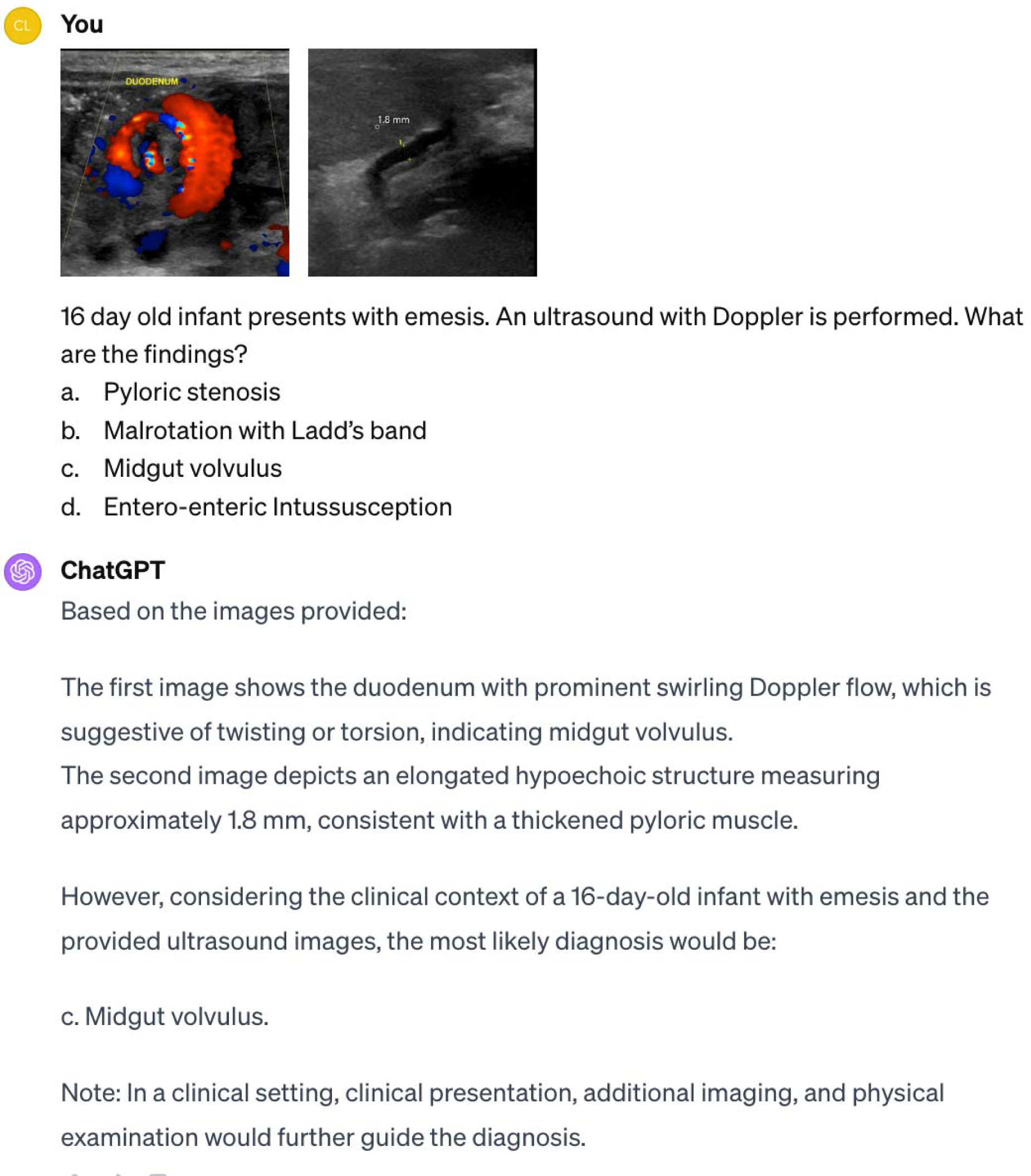
ChatGPT answers correctly a Pediatric Radiology higher-order question with two images, and recognizes the imaging findings on the images. However, while it recognizes the signs of midgut volvulus on the first image, it makes a factually incorrect assertion of identifying the thickness of a single pyloric muscular wall on a transverse image measuring 1.8 mm as thickened (threshold >3mm). However, using the clinical context in the question along with the “whirlpool sign” in the first US image, it correctly chooses Option C – Midgut volvulus.

**Figure 4:**
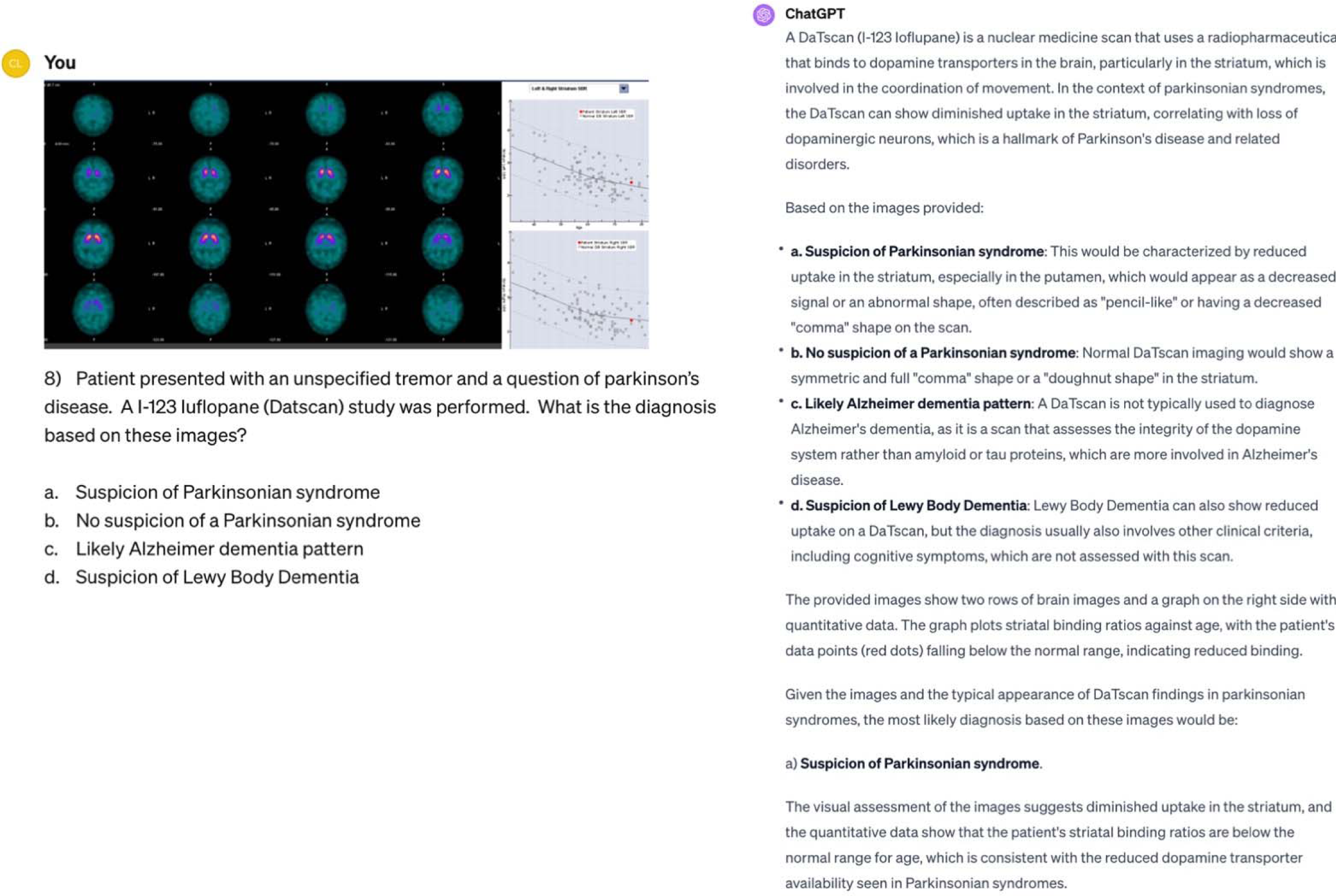
ChatGPT’s answer to a higher-order question in Nuclear Radiology in examining a DaTscan images along with the accompanying graphs. ChatGPT chooses the incorrect answer (Option A) instead of Option B: No suspicion of a Parkinsonian syndrome. It analyzes the images incorrectly suggesting diminished uptake in the striatum, and striatal binding ratios as below normal range for age (the images however show normal striatal uptake with normal binding ratios)

### Performance by Domains

Among the subspecialty domains, there was overall no significant difference in performance (P=.76). However, ChatGPT performed best in Cardiovascular imaging with 80% (8 out of 10) while its performance in Thoracic imaging was 30% (3 out of 10) without being significantly different (P=.07).

**Table 2:** ChatGPT performance for the second run from November 30-December 10 overall and sub-stratified across question types domains, modality, difficulty. ^ P value represent performance comparison between lower-order and higher-order questions * P values represent pairwise comparisons to performance on lower-order thinking using Fisher’s exact test. Please note that no pairwise comparisons were performed between the clinical domains since P-value from Fisher’s exact test did not show significant difference between the domains.

| Question Type | No. of Questions | No. of Correct Responses | P value |
| --- | --- | --- | --- |
| All questions | 110 | 60 (55%) |  |
| Question Type |  |  |  |
| Lower-order thinking | 41 | 23 (57%) |  |
| Higher-order thinking | 69 | 37 (54%) | .85^ |
| Probable diagnosis | 16 | 10 (63%) | .77* |
| Providing clinical management and follow-up imaging recommendations | 13 | 8 (62%) | .76* |
| Synthesizing information from different sequences | 18 | 12 (67%) | .40* |
| Pathophysiology or Structure affected | 22 | 7 (32%) | .12* |
| Domain |  |  | .17 |
| Breast | 10 | 2 (20%) |  |
| Cardiovascular | 10 | 7 (70%) |  |
| Gastrointestinal | 10 | 7 (70%) |  |
| Genitourinary | 10 | 4 (40%) |  |
| Interventional | 10 | 7 (70%) |  |
| Musculoskeletal | 10 | 4 (40%) |  |
| Neuroradiology | 10 | 7 (70%) |  |
| Nuclear Radiology | 10 | 4 (40%) |  |
| Pediatric Radiology | 10 | 7 (70%) |  |
| Thoracic Imaging | 10 | 4 (40%) |  |
| Ultrasound | 10 | 7 (70%) |  |
| Modality |  |  | .42 |
| Radiography and Mammography | 18 | 8 (44%) |  |
| CT | 34 | 18 (53%) |  |
| MRI | 24 | 17 (71%) |  |
| Ultrasound | 20 | 10 (50%) |  |
| Other (Fluoro, DSA, Nuclear medicine, PET) | 14 | 7 (50%) |  |
| Difficulty by Radiologists |  |  | .25 |
| Low (1-5) | 60 | 36 (60%) |  |
| High (6-10) | 50 | 24 (48%) |  |
| Difficulty by Trainees |  |  | .44 |
| Low (1-5) | 56 | 33 (59%) |  |
| High (6-10) | 54 | 27 (50%) |  |

### Performance by Modalities

There was no significant difference in performance when questions were divided by modalities (P=.66) However, performance in questions involving MRI (67%, 16/24) was the best while it answered 47% (16/34) correctly when the images were CT scans.

### Performance by Difficulty

ChatGPT performed better on questions rated on a difficulty scale of 1-5 assigned by both radiologists (62%, 37/60) and radiologists-in-training (63%, 35/56) when compared with higher-order questions (48%, 24/50 and 48%, 26/54 respectively) but was not significantly different (P=.17).

There was substantial agreement in grading the difficulty between the radiologists and the radiologists-in-training [Kappa - .63 (95% CI - .49 - .78)]

### Confidence level and qualitative observations

All the questions were deemed appropriate for a board style examination by two 3^rd^ year radiologists-in-training who were preparing for boards and were not involved in composing the questions.

In the majority of questions (89%; 98/110), ChatGPT was confident or very confident in its answers. In the remainder of the questions (12/110), it answered 75% (9/12) incorrectly. In 39% (43/110), it incorrectly or could not recognize (5/43) the imaging finding but correctly answered 12% of the questions correctly (5/43) by using the text-based portion of the question. There was a significant difference in confidence level between correct and incorrect answers (Mean Likert score 4.7 vs 4.1 respectively; P=.01). However, even in the questions it answered incorrectly, it used confident or very confident terminologies (82%; 40/49).

Factually incorrect assertions include identifying the thickness of a single pyloric muscular wall on a transverse image measuring 1.8 mm as thickened (Figure 3) in a question it also correctly identified the whirlpool sign of midgut volvulus in a vomiting newborn. It also erroneously claimed that the diagnostic criteria for polycystic ovarian syndrome was >15 follicles per ovary.

In 6% (7/110) questions ChatGPT had difficulty in choosing between two options, choosing incorrectly 4/7 times.

### Assessing reproducibility

In the second run of ChatGPT (performed between November 30 – December 10) overall performance was 54% (59 out of 110). Applying the correction-for-guessing formula, performance was 38% (42 out of 110) respectively. Cohen’s Kappa for agreement in the two runs was .73 (95% CI - .60 - .85) indicating substantial agreement.

While performance was grossly similar across the different subcategories when compared between the two runs, performance in Breast Radiology worsened on the second run (2/10 vs 5/10; p= .36) while performance improved in Interventional and Neuroradiology (7/10 vs 5/10; p=.65) (Table 3). While there was substantial agreement between the two runs, there were 7/110 questions that ChatGPT answered correctly in the second run after answering incorrectly in the first run. Two out of these questions, it was not confident in its reasoning. All of these were higher order questions. Meanwhile, it answered 8/110 questions incorrectly in the second run, after answering it correctly in the first run. These included two lower-order questions and the remaining higher order questions and was confident in its reasoning both times (7/8). There was no significant difference in difficulty between the questions (6/15 questions with difficulty 1-5; P=.42)

**Table 3:** Comparing ChatGPT performance across both runs overall and sub-stratified across question types domains, modality, difficulty.

| Question Type | No. of Questions | No. of Correct Responses |  | P value |
| --- | --- | --- | --- | --- |
|  |  | 1<sup>st</sup> Run | 2<sup>nd</sup> Run |  |
| All questions | 110 | 61 (55%) | 60 (55%) | >.99 |
| Question Type |  |  |  |  |
| Lower-order thinking | 41 | 25 (61%) | 23 (57%) | .83 |
| Higher-order thinking | 69 | 36 (52%) | 37 (54%) | >.99 |
| Probable diagnosis | 16 | 8 (50%) | 10 (63%) | .72 |
| Providing clinical management and follow-up imaging recommendations | 13 | 6 (46%) | 8 (62%) | .70 |
| Synthesizing information from different sequences | 18 | 12 (67%) | 12 (67%) | >.99 |
| Pathophysiology or Structure affected | 22 | 10 (45%) | 7 (32%) | .54 |
| Domain |  |  |  |  |
| Breast | 10 | 5 (50%) | 2 (20%) | .36 |
| Cardiovascular | 10 | 8 (80%) | 7 (70%) | >.99 |
| Gastrointestinal | 10 | 7 (70%) | 7 (70%) | >.99 |
| Genitourinary | 10 | 5 (50%) | 4 (40%) | >.99 |
| Interventional | 10 | 5 (50%) | 7 (70%) | .65 |
| Musculoskeletal | 10 | 5 (50%) | 4 (40%) | >.99 |
| Neuroradiology | 10 | 5 (50%) | 7 (70%) | .65 |
| Nuclear Radiology | 10 | 5 (50%) | 4 (40%) | >.99 |
| Pediatric Radiology | 10 | 7 (70%) | 7 (70%) | >.99 |
| Thoracic Imaging | 10 | 3 (30%) | 4 (40%) | >.99 |
| Ultrasound | 10 | 6 (60%) | 7 (70%) | >.99 |
| Modality |  |  |  |  |
| Radiography and Mammography | 18 | 10 (56%) | 8 (44%) | .74 |
| CT | 34 | 16 (47%) | 18 (53%) | .81 |
| MRI | 24 | 16 (67%) | 17 (71%) | >.99 |
| Ultrasound | 20 | 12 (60%) | 10 (50%) | .75 |
| Other (Fluoro, DSA) | 14 | 7 (50%) | 7 (50%) | >.99 |
| Difficulty by Radiologists |  |  |  |  |
| Low (1-5) | 60 | 37 (62%) | 36 (60%) | >.99 |
| High (6-10) | 50 | 24 (48%) | 24 (48%) | >.99 |
| Difficulty by Trainees |  |  |  |  |
| Low (1-5) | 56 | 35 (63%) | 33 (59%) | .85 |
| High (6-10) | 54 | 26 (48%) | 27 (50%) | >.99 |

Figure 5 shows a question it answered correctly in the first run, but chose wrong in the second run.

**Figure 5:**
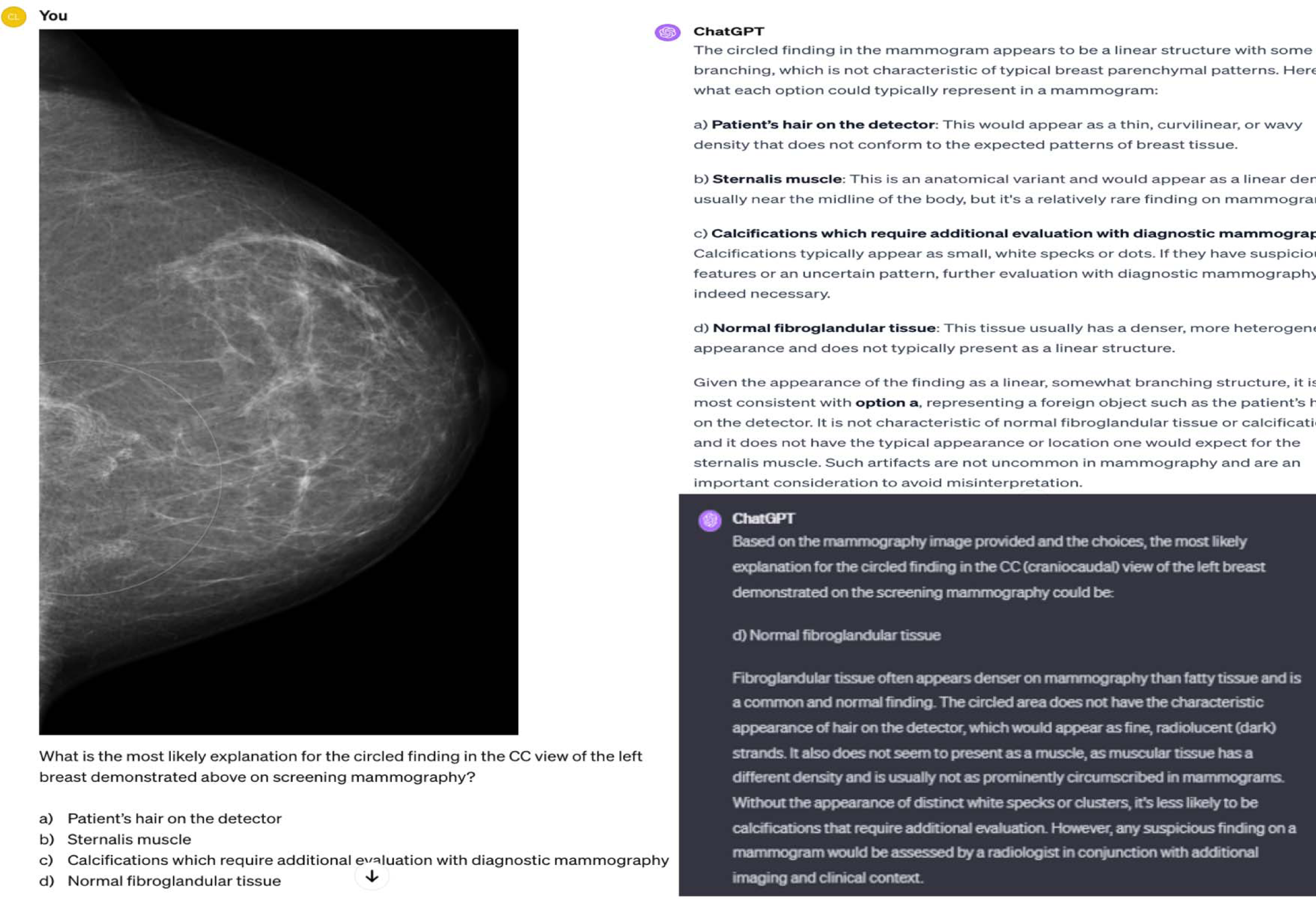
In a lower-order question from Breast Imaging ChatGPT 4 perfectly identifies the hair artifact on the first run on the mammogram images (Top Right) and chooses the correct Option A. In the second run (Bottom right) it however does not recognize the finding on the image and hence chooses Option D – Normal fibroglandular tissue incorrectly.

## DISCUSSION

In our study assessing the performance of ChatGPT 4 in a radiology board-style examination with questions containing images, overall performance was 55% out of 110 multiple-choice questions, indicating that it fails to clear the exam (considering a passing score of 70%). While it performed better on lower-order questions (61%) as compared to higher-order (52%), results were not significantly different (P=.43). Amongst the different higher-order categories it did better on synthesizing information from different imaging modalities/sequences (67%) as compared to the other categories (probable diagnosis (50%), best clinical management/imaging follow-up (46%) and pathophysiology (45%).

ChatGPT was confident in reasoning even when incorrect (89%) even though there was a different in confidence between correct and incorrect answers (Mean Likert score 4.7 vs 4.1; P=.01). In assessing reproducibility, there was a substantial agreement between two runs (Kappa - .66 (.60-.85)), even though it changed answers in 14% of questions (15/110). This performance is promising when it comes to the recently unveiled multimodal capabilities of an LLM without any medical or radiology specific pre-training.

Previously published studies about ChatGPT performance in medical exams including radiology board style examinations were limited by the capabilities of the model at that time in being constrained to only text-based prompts. This limited accurate simulation of a radiology board examination which is image-rich. Ours is the first of its kind study to closely approximate a radiology board style examination with questions containing images and text. Additionally, we designed the questions using the ABR Core examination blueprints incorporating image-rich questions from the eleven clinical components of radiology that is tested in the examination. As expected due to our questions containing images and text, performance was worse (55%), as compared to ChatGPT 3.5 (69%) and ChatGPT 4(81%) on text based questions only. Moreover, while performance was better on lower-order vs higher-order questions, and similar to prior studies, the results weren’t significantly different. Additionally, in our study there was a significant difference in confidence between correct and incorrect answers (P=.01), even though it answered incorrect questions with confident language (Mean Likert score – 4.1). Bhayana et al. showed that ChatGPT answered 100% questions using confident/highly confident language (as compared to 89% in our study).

While ChatGPT might not have been able to “pass” the examination, the capabilities of a model not pre-trained or without domain specific knowledge is intriguing. Its relative ease in recognizing imaging findings and answering complex questions, synthesizing information from both images as well as the clinical information provided shows great potential. Additionally, it successfully simulated human exam taking behavior as in a few cases (5/110) where while it failed to recognize the imaging finding or incorrectly recognized the finding, it managed to answer the correctly by reasoning out the text based question stem and answer choices, eliminating the probable wrong options.

However, before radiologists-in-training can use ChatGPT as a tool to analyze potential difficult or challenging images or use it as a companion in learning radiology, our study revealed several limitations. The primary limitation was lack of reliability as in a second run using the same version of ChatGPT Plus but from a different account, 14% (15/110) questions had a different response, in spite of otherwise substantial agreement (Kappa - .73 (95% CI - .60 - .85), as compared to the initial run. This calls into question its reliability and the process behind how it analyzes information. This was similar to reported results from Bhayana et al^13^. using a text-based exam between ChatGPT 3.5 and 4. However, our results are even more concerning since we used the same ChatGPT 4 model for both simulations. Even after excluding the cases (7/110) where ChatGPT showed uncertainty in having to choose between two options ChatGPT used confident terminologies (89%; 98/110), “hallucinations” even when answering incorrectly (82%; 40/49), which makes it highly unreliable (Mean Likert score – 4.1). This also included factual inaccuracies such as discrepancies in the standardized metric for pyloric wall thickening and criteria for polycystic ovarian syndrome diagnosis. These errors provide a cautionary tale before we employ LLMs as the primary knowledge source. Additionally, while it successfully answered several higher-order questions correctly, it failed to answer simple lower-order questions such as identifying a renal cyst on ultrasound, the Fleischner criteria, recognizing pulmonary embolism on perfusion scans.

Our study had several limitations. Firstly, questions were not official ABR or Canadian Royal College questions. Secondly, the real-world test includes a combination of both image-based and text questions, but we included image-based questions only as ChatGPT’s performance on text-based questions has already been comprehensively studied. Thirdly, only still images (upto 4) per question was prompted to ChatGPT 4, with no scrollable images or videos as part of the questions, due to current technical limitations of ChatGPT. Moreover, the passing grade is only an approximation, and we did not use criterion-based scoring that the ABR uses. Finally, while we developed the questions using the ABR core blueprint, the number of questions were relatively low in the exploratory study and the subgroup analysis was underpowered.

In conclusion, our study shows promising performance of the multimodal capability of ChatGPT Plus in a radiology board-style examination containing image-rich questions, even though it fell well short of a probable passing mark. While performance was not significantly different among different question categories, it performed better on lower-order and lower difficulty questions. While promising, our study shows that the tool is not ready to be used by radiologists or radiologists-in-training for learning or practicing since it repeats previously reported limitations, including confidence in answering incorrectly, factual inaccuracies and lack of reproducibility even in the same software version. However, recognizing the strength and limitations of LLMs like ChatGPT is crucial for radiologists to be gate-keepers in using novel AI-enabled technologies in their field.

## Data Availability

All data produced in the present work are contained in the manuscript

## Notes

### Competing Interest Statement

The authors have declared no competing interest.

### Funding Statement

The study did not receive any funding.

